# Performance of Hb HemoCue machine compared to automated hematology analyzer for hemoglobin measurements among adult patients at Kilimanjaro Christian Medical Centre

**DOI:** 10.1101/2024.12.07.24318646

**Authors:** Nancy A. Kassam, Goodluck A. Mwanga, Elia L. Yusuph, Elda M. Maundi, Mose Josephat, Neema B. Kulaya, Daniel B. Lasway, Zacharia L. Laizer, Goodluck G. Ndossy, James S. Kimaro, Arnold Ndaro

## Abstract

**Background:** Automated hematology analyzers offer precise hemoglobin measurements, but are expensive and impractical for field, point of care, primary care and remote settings use. The portable and cost-effective Hemocue device provides an alternative. Comparing their accuracies is crucial to prevent diagnostic discrepancies and misdiagnoses. This study aimed to determine the accuracy of Hb HemoCue machine by comparing its performance to automated analyzer at KCMC clinical laboratory where both equipment are used.

**Methods:** A cross-sectional study was conducted at Kilimanjaro Christian Medical Centre (KCMC) Clinical Laboratory among adult patients whose hemoglobin concentrations were measured in May to June 2024. Hemoglobin levels were estimated using two distinct methods: the Hb HemoCue machine and repeatedly tested using an automated hematology analyzer.

**Results:** Hemoglobin (Hb) concentration values obtained from the HemoCue machine and the automated analyzer, had a mean difference of 0.001 g/dl (95% Cl: -0.036 to 0.038), t value of 0.062, and a *p*-value of 0.95, indicating a non-statistically significant differences between the two measurement methods. The Bland-Altman plot analysis indicated that the mean difference (bias) between the two methods was 0.0012 g/dL, and the limits of agreement ranged from - 0.481 to 0.482 g/dL, suggesting that the HemoCue machine tends to slightly overestimate Hb values compared to the automated hematology analyzer. The Pearson correlation coefficient for the Hb concentrations measured using HemoCue and automated analyzer was 0.995, indicating a very strong positive correlation. Receiver operating characteristics (ROC) curve showed that the area under the curve (AUC) for analyzer and HemoCue was 1.000 indicating that both methods have good diagnostic accuracy of measuring Hb concentrations.

**Conclusion:** The study revealed strong agreement between HemoCue and automated hematology analyzer for measuring hemoglobin concentrations. Both methods demonstrated high diagnostic accuracy suitable for clinical use. Although HemoCue slightly overestimated hemoglobin, this difference was deemed insignificant. The study endorses HemoCue as a reliable tool for hemoglobin concentration measurement alongside automated analyzers.

## Background

The most trustworthy metric for screening people for anemia and gauging the effectiveness of medical and dietary interventions is hemoglobin (Hb) measurements. Hemoglobin levels are one of the most accurate markers of anemia and are frequently used to identify anemic people and monitor how well interventions are working [1,2].

The World Health Organization color scale, Sahli’s method, HemoCue, and clinical examination for pallor are frequently used techniques to estimate hemoglobin in a community, and primary healthcare facilities [3,4]. However, these techniques have a number of drawbacks, including poor accuracy, complexity, and high expense [4,5]. Accurate quantitative diagnostic tests can verify the diagnosis of anemia, nevertheless, highly accurate techniques either depend on constant electricity supply, use costly or toxic chemicals and consumables, or require constant quality control and needs to be operated by a trained personnel [1]. In most cases, these are not appropriate for use in the majority of point of care and primary health-care settings with limited resources [1,2].

In clinical laboratories, automated hematology analyzers are typically used to measure Hb concentrations. These instruments are highly accurate and dependable, but they are also costly and impractical for field, point of care and primary healthcare use [2,6]. On the other hand, the HemoCue device has been widely used in field settings with limited resources due to its portability, ease of use, and reasonable cost. Additionally, the HemoCue device gives an instant numerical Hb value with just a tiny drop of capillary or venous blood [1].

At Kilimanjaro Christian Medical Centre (KCMC) Clinical Laboratory, HemoCue machine is used alongside the automated hematology analyser. To save costs of operation, the Hb HemoCue machine is used when the test request needs only hemoglobin levels to be measured while the automated analyser is used when the test request needs a broader hematological profile to be analyzed. It is crucial to assess the accuracy of HemoCue compared to that of automated hematology analyzers in hemoglobin estimation. Understanding the differences between these two methods is essential for preventing misdiagnoses and understand if there are significant results discrepancies between Hb values when measured using the two different equipment.

In Tanzania, there is limited published data from studies comparing the performance of HemoCue and automated hematology analyzers among patients undergoing hemoglobin testing in clinical laboratories, despite the HemoCue machine’s widespread use. This study compared Hb HemoCue performance to that of an automated hematology analyzers among adult patients undergoing hemoglobin concentrations testing at KCMC clinical laboratory.

## Materials and Methods

### Study site

This study was conducted at KCMC Clinical Laboratory, which is a well-equipped medical facility with the necessary infrastructure and expertise to conduct hemoglobin tests using both HemoCue and automated analyzer. Kilimanjaro Christian Medical Centre (KCMC) is a referral zonal hospital situated in Moshi, a town located within Kilimanjaro Region in Tanzania, East Africa. KCMC offers a wide range of inpatient and outpatient medical services. It serves over 15 million people in the northern zone of the country and beyond attending outpatients and 500-800 inpatients per day with 630 bed capacity. The clinical laboratory is one of the hospital’s department which offers a wide range of diagnostic tests including hematological tests.

### Study design and sampling procedures

A cross-sectional study was conducted from 1^st^ May to 30^th^ June 2024 to compare hemoglobin concentrations and accuracy obtained using HemoCue machine and an automated hematology analyzer among adult patients. A convenient sampling technique was used to obtain participants whose blood samples were used in this study. Individuals aged 18 years and above, who came to KCMC Clinical laboratory with blood test requests for full blood count and consented to participate in this study were included. A total of 170 individuals were enrolled in this study and in total, 170 blood samples were collected and analysed for hemoglobin concentrations.

### Ethical approval

Ethical approval for this study was obtained from the Kilimanjaro Christian Medical University College, College Research Ethics and Review Committee (KCMUCo-CRERC) (certificate number UG 105/2024). Written informed consent forms were given to potential participants, and those who gave consent were included in the study. Participants’ information were only accessed by the research team and only identification numbers were used in data compilation to ensure confidentiality.

### Data collection

Permission for data collection was sought from KCMC Hospital and Clinical Laboratory management. A simple data collection log was used to obtain participants’ information including a unique identifier, age and gender and Hb concentration.

#### Collection of venous blood

Blood collection procedure was well explained to the patient followed by the process of blood collection. A tourniquet was applied just above the venipuncture site and sterile alcohol swabs used to clean the area of venipuncture. Blood was drawn from an antecubital, dorsal metacarpal or great saphenous vein into an EDTA vacutainer using a sterile syringe. Blood sample tubes were labeled with patient information including the study identification number and sent to the clinical laboratory for analysis.

#### Laboratory procedures

The collected samples were tested for hemoglobin concentration (Hb) using both automated hematology analyzer (AHA) (Mindray_BC-5380, Shenzhen Mindray Bio-Medical Electronics Co. LTD) followed by Hb Hemocue machine (HemoCue 201+, HemoCue AB, Sweden). The results of hemoglobin values obtained from both the Hemocue machine and the automated hematology analyzer for each patient were recorded in the data collection log.

For quality control, daily maintenance and daily quality control of automated hematology analyzer were performed. Routine thorough clean-up of equipment was performed as required in the daily maintenance standard operating procedure. Daily quality control was performed by analyzing three quality control samples (BC-5C Hematology Control, Mindray) which measures high, normal and low hematological parameters.

### Statistical analysis

Data entries were double checked and final database was cleaned and analysed using Statistical Package for Social Sciences (SPSS) software version 25. The mean of differences (bias), standard deviation of differences (SD) and limits of agreement between the HemoCue and the automated hematology analyzers readings were calculated using Bland-Altman model. Pearson correlation analysis was performed and correlation coefficient (r) was calculated. Paired t-tests tests were used to compare the difference between the mean Hb concentrations values estimated by HemoCue and the automated hematology analyzer. The automated hematology analysers was considered as the reference method against which the accuracy of the HemoCue was determined. Frequency distributions of categorical variables such as gender and age groups were determined. Results were presented in figures, and summarized in tables, and significant differences or correlations were determined at *p* < 0.05.

### Variables

The dependent variables included hemoglobin concentrations and independent variables included method of estimation of hemoglobin concentrations (Hemocue and automated hematology analyzer) and demographic variables (age and gender). The hemoglobin concentrations were presented as continuous variables and age and gender were treated as categorical variables.

## Results

### Characteristics of the study population

A total of 170 adults of 18 years and above participated in this study. All 170 participants were tested for hemoglobin concentrations using both the Hb HemoCue and automated hematology analyzer. Distribution by gender was nearly equal, with 52.0% being female individuals. The participants’ mean age was 52.2 years and standard deviation (SD) was 20.05 years (Table 1).

**Table 1:** Characteristics of the study population (N=170)

| Variable | N | % |
| --- | --- | --- |
| **Age (years)** |  |  |
| 18-30 | 36 | 21.2 |
| 31-40 | 24 | 14.1 |
| 41-51 | 8 | 4.7 |
| >51 | 102 | 60.0 |
| **Mean (SD)** | **52.2(20.05)** |  |
| **Sex** |  |  |
| Male | 81 | 47.6 |
| Female | 89 | 52.4 |

**Table 1:** Comparison of the Hb concentrations measured by two methods for hemoglobin estimation, Hemocue and automated hematology analyzer (AHA), N=170.

|  | Minimum<br>conc. | Maximum<br>conc. | Mean | Std. Deviation | Std. Error<br>Mean |
| --- | --- | --- | --- | --- | --- |
| AHA (g/dl) | 5.8 | 17.8 | 11.605 | 2.3974 | 0.1839 |
| Hemocue (g/dl) | 5.7 | 17.9 | 11.606 | 2.4613 | 0.1888 |

STD - standard, g/dl - gram per deciliter, conc – concentration, AHA – automated hematology analyser

### Comparison of the Hb concentrations measured by HemoCue and AHA

Descriptive statistics were calculated for Hb concentrations from both HemoCue and the automated hematology analyzer. For 170 readings, the mean Hb concentration value for HemoCue was 11.606 g/dl with a standard deviation of 2.461 g/dl and standard error of mean was 0.1888, while for the same number of readings, the mean Hb concentration value for analyzer was 11.605 g/dl with a standard deviation of 2.397 g/dl and standard error of mean was 0.1839 (Table 2). The mean difference for the two equipment measurements was 0.001g/dL.

The Pearson correlation coefficient for the HB concentrations measured using HemoCue machine and automated hematology analyzer was 0.995 and this correlation was statistically significant p<0. 001.

A pair sample t-test was conducted to compare the difference in Hb concentration values obtained from HemoCue machine and the automated hematology analyzer. The result showed a mean difference of 0.001 g/dl (95% Cl: -0.036 to 0.038). The t value was 0.062, with a *p*-value of 0.95, indicating that the differences between the two Hb concentration measurement methods are not statistically significant.

### Bland-Altman comparison of the hemoglobin concentrations measurements performed in venous blood samples by AHA and HemoCue machine

Data in **Figure 1** below represents the differences (automated hematology analyzer–HemoCue) for the average Hb results. The middle solid line represent the mean difference between the measurements (bias), while the upper and lower lines indicate the 95% limits of agreement between methods. The mean difference (bias) was 0.0012g/dl, with the limit of agreement ranging from -0.481 to 0.482g/dl. This suggest that HemoCue tends to slightly overestimate Hb values compared to the analyzer, but the differences are generally small and within acceptable limits.

**Figure 1:**
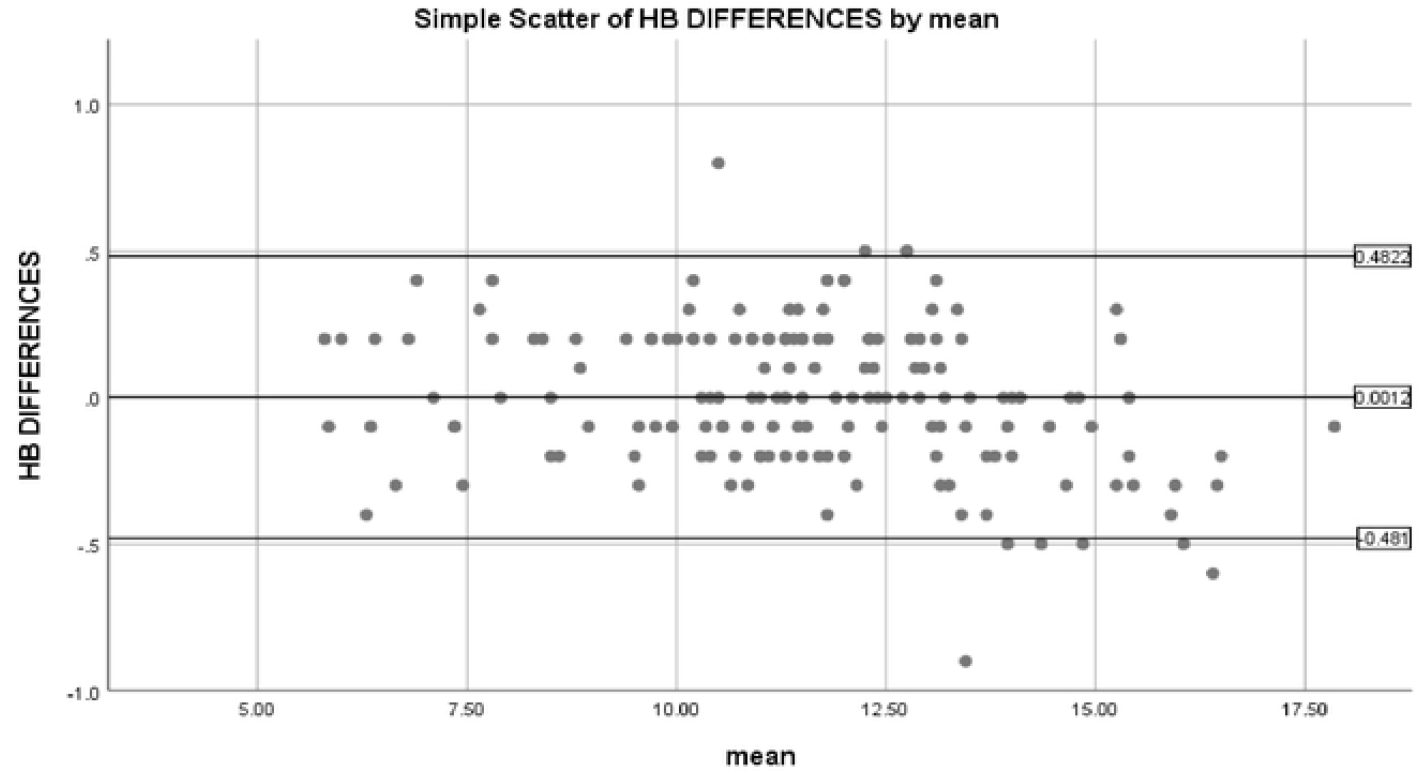
Bland-Altman comparison of the hemoglobin concentration measurements performed in venous blood samples by hematology automated analyzer and Hb HemoCue.

### Accuracy of HemoCue in hemoglobin estimation as compared to automated hematology analyzer

In **Figure 2**, the area under the curve (AUC) for automated hematology analyzer and HemoCue was 1.000, indicating that both methods have good diagnostic accuracy. These finding support the use of both methods for Hb measurements in clinical settings.

**Figure 2:**
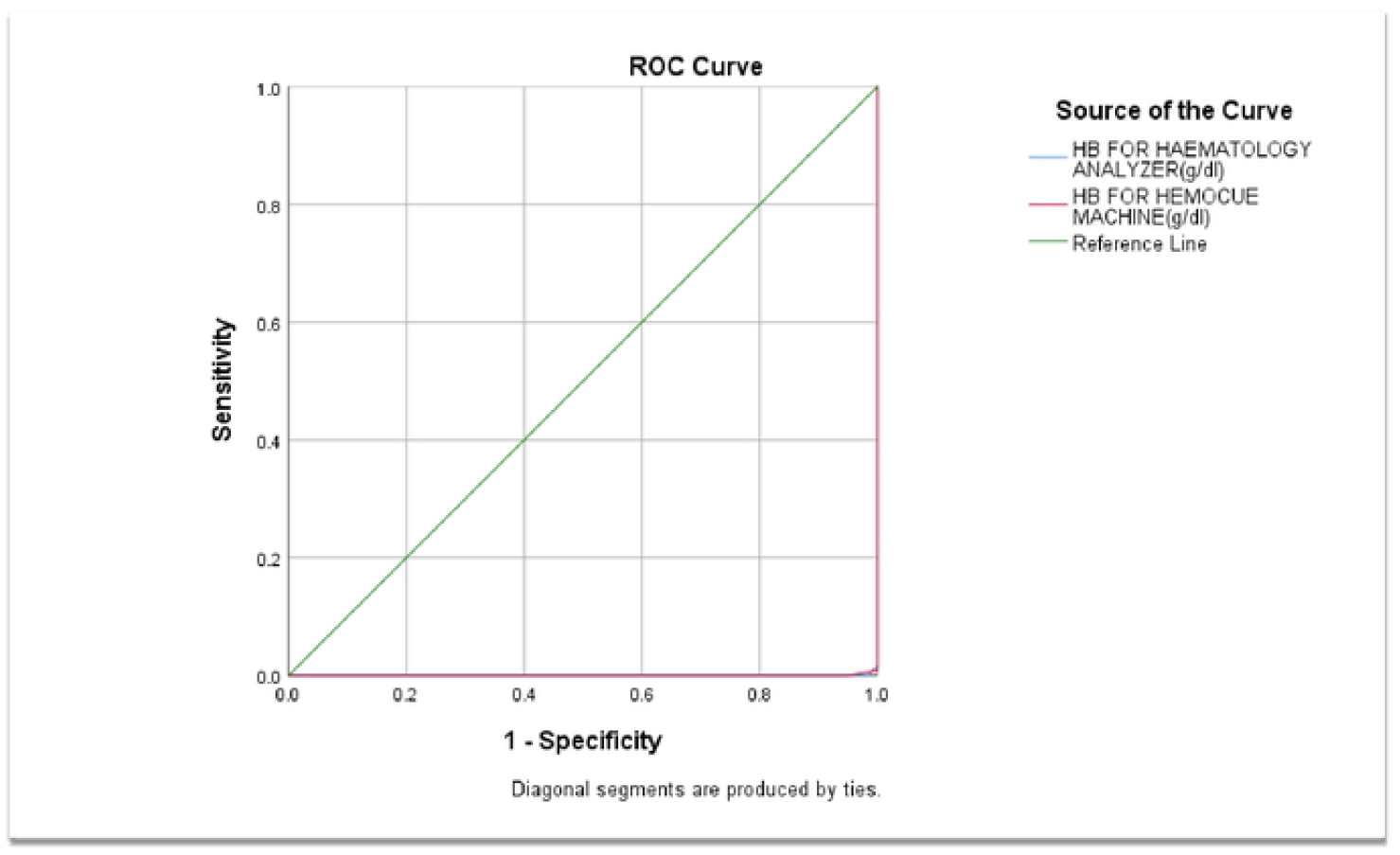
ROC curve comparing hemoglobin concentrations measured using HemoCue machine to those measured using automated hematology analyzer.

## Discussion

The results of this study have important implications for clinical practice, particularly in settings where different methods are used interchangeably and in low resource and point of care settings where simple yet reliable methods are needed for appropriate diagnosis of anaemia.

While the results of this study show that, the HemoCue machine slightly overestimates Hb concentration values (mean difference 0.001g/dL), this overestimation is minimal and may not be clinically relevant in the current study setting. Another study reported a mean difference of 0.2 g/dL, for Hb concentration measured by HemoCue compared to an automated analyser Sysmex KN21TM, reinforcing the observation that while the HemoCue is reliable, it systematically reads slightly higher than automated analyzers [7].

The result of this study show, both the HemoCue machine and the automated hematology analyzer provide reliable and consistent Hb measurements. Although there is a slight variation in Hb concentrations measured by the two methods, the differences are within clinically acceptable limits. The Pearson’s correlation coefficient approached 1 (r=0.995; *p*<0.001), indicating a significantly strong positive correlation between the Hb concentration values measured by the two equipment. Our study’s findings are further supported by the paired sample t-test results which revealed a lack of statistically significant difference between the Hb values obtained from the HemoCue machine and the automated hematology analyzer, with a mean difference of 0.001 g/dl (95% CI: -0.036 to 0.038), t value of 0.062, and a p-value of 0.95.

Concurrent with the current study’s findings, other studies also showed that the Hb concentration results measured using HemoCue machine significantly correlate to the Hb concentration results measured by automated analysers as the standards [7–10], supporting the reliability and accuracy of HemoCue in the places where automated analysers are impractical.

Contrary to the current study’s findings, other studies reported a lack of or weak correlation between hemoglobin concentrations when measured by HemoCue and automated analyser, probably because they compared certain specific populations or compared different types of samples e.g. capillary versus venous blood [11,12].

The Bland-Altman plot analysis indicated that the mean difference (bias) between the two methods was 0.0012 g/dL, and the limits of agreement ranged from -0.481 to 0.482 g/dL. This analysis highlights the variability between the two measurement techniques and emphasizes that, while the mean difference is minimal, there is a substantial range of agreement which may need to be given consideration during clinical interpretation of results depending on how critical the case is. The differences are generally small as they are within 0.5 limits of agreement which is within the acceptable limits (±1) [9,13]. These findings concurred with what Salmond et al had reported [13]. Other studies carried out in Khartoum and Khammouane Province in Lao among pregnant women and children respectively demonstrated rather higher and therefore poor limits of agreement (bias of 1.17 LoA ± 1.57 and bias of 6.1 g/L and LoA −11.5 g/L to 23.7 g/L respectively) for venous blood samples when Hb was estimated using HemoCue and automated analyser [9,14]. Unlike the current study, Adam et al compared HemoCue and Sysmex automated analyser and the different results may also be subjected to other factors such as the competence of the person performing the test and the performance of the Sysmex analyser in comparison to that of HemoCue. Furthermore, it is suggested that capillary blood sampling may result in pre-analytical errors which can influence results for point of care devices [15].

The accuracy of HemoCue in hemoglobin estimation as compared to automated hematology analyzer under ROC curve showed the area under the curve (AUC) for analyzer as a standard and HemoCue to be 1.000, indicating that Hb HemoCue machine has good diagnostic accuracy for measuring anemia. However, clinicians should consider cross-verifying critical Hb values with an automated analyzer when possible. The current study’s findings support the use of both methods for Hb measurement using venous blood in clinical settings but other studies done in Gambia shows HemoCue proved superior to Aptus analyzer in accuracy of measuring non-anemia, with an AUC of 0.933 compared to 0.799, respectively [16], while another study conducted among women of reproductive age in Peru reported the automated analyser to be superior to HemoCue (AUC 0.82 and 0.71 respectively) [17]. These discrepancies are probably because in the current study, a lot of factors were kept the same. For instances, only venous blood samples were used for both analysers and the same person operated the equipment during measurements. Additionally, the tests were performed by qualified laboratory technologists in an accredited laboratory, which implies that there are high performance standards.

Policy-wise, while HemoCue is valuable for its convenience and rapid results, automated analyzers should be preferred for detailed diagnostics and critical decisions. Training healthcare providers to recognize and interpret these potential differences can enhance patient safety and care quality. The use of point of care devices, however, should fulfill some basic criteria, including economic, clinical, and regulatory issues; appropriate training of the users and knowledge of test requirements, performance, limitations, and potential interferences; the use of venous and arterial sampling, when possible; and a rigorous quality assessment, which should be under the responsibility of laboratory professionals [18,19].

## Limitations

This study compared Hb concentrations and accuracy of HemoCue and automated hematology analyser using only venous blood. Further studies can be conducted in this setting to compare the performance of the two equipment and the potential for results variations when capillary blood is used compared to when venous blood is used. This study predominantly focused on adults aged above 18 years and venous blood samples. Future research should explore how age, sex, and different clinical conditions and type of sample (capillary versus venous blood) could influence Hb measurements obtained from both HemoCue and the automated hematology analyzer.

## Conclusion and Recommendation

The study concludes that there is a high level of agreement between the HemoCue and automated hematology analyzer, Mindray BC-5380 in measuring hemoglobin concentrations. Both methods demonstrate excellent diagnostic accuracy and can be reliably used in clinical settings for Hb estimation. The slight overestimation by the HemoCue is negligible, and thus, it is a valid tool for Hb measurement alongside the automated hematology analyzer. This study’s findings therefore recommended HemoCue for use as a point of care device for determining hemoglobin in resource limited areas as well as critical care areas of health facilities.

## Data Availability

All data is available, and will be provided after acceptance for publication.

